# Using Multimodal Data Collection System as a Research Tool in the Major Depressive Disorder Analysis: a cross-sectional study protocol

**DOI:** 10.1101/2024.07.21.24310061

**Authors:** Hongbo Li, Yifu Ji, Linxiang Xu, Jiaoyun Yang, Yang Du, Min Hu, Ning An

**Affiliations:** Anhui Province Key Laboratory of Affective Computing and Advanced Intelligent Machine, Hefei University of Technology Hefei 230601, China; Affiliated Psychological Hospital of Anhui Medical University, Hefei, China; Anhui Mental Health Center, Hefei, China; the Fourth People’s Hospital of Hefei,Hefei 230032,China; National Smart Eldercare International Science and Technology Cooperation Base, Hefei University of Technology Hefei 230601; Intelligent Interconnected Systems Laboratory of Anhui Province (Hefei Universityof Technology), Hefei, 230009, China

## Abstract

**Introduction:** Previous studies have established that depressive syndromes can be detected using machine learning methods, with multimodal data being essential. Multimodal data facilitates the extraction of characteristics such as gaze tracking, a reliable depression indicator. Our study employs high-quality video and other multimodal data from patients diagnosed with depression. Our study uses a multimodal data collection system (MDC) to understand the complex indicators of depression.

**Objective:** This paper outlines our protocol for deploying a multimodal data collection system within an In-Person Clinical Assessment environment. The system gathers high-definition videos, real-time vital signs, and voice recordings for future extraction of critical information such as eye gaze patterns. We aim to scale our model to provide portable depression risk analyses, facilitating timely intervention and encouraging patients to seek professional assistance.

**Methods and Analysis:** We have conducted sessions with 70 participants diagnosed with depression. Each participant undergoes DSM-5 interviews and engages with our multimodal data collection system. Participants respond to five on-screen scales while being recorded. To our knowledge, no other protocol has combined multimodal data collection and various stimuli in depression data collection.

**Ethics and Dissemination:** Ethical approval was provided by the National Health Commission of the PRC, Hefei Fourth People’s Hospital Ethics Committee (HSY-IRB-YJ-YYYX-JYF001). Results will be published in a peer-reviewed journal and presented at academic conferences.

## INTRODUCTION

### Background

According to the World Health Organization, 3.8% of the global population, approximately 280 million people, suffer from depression, with over 75% in middle- and low-income countries not receiving adequate treatment (1). Depression is a leading cause of suicide, especially among adolescents, yet its precise causes remain unclear (2). Diagnostic methods, primarily based on DSM-5 criteria, typically require multiple healthcare professionals to reduce subjectivity (3). During interviews, patients may withhold information, complicating diagnosis.

Deep learning has shown promise in early depression detection and treatment. University of Edinburgh research suggests behavioral indicators, such as facial and eye movements, offer a cost-effective diagnostic alternative (3). Studies using facial electromyography and movement ratings during emotional content exposure achieved 79.17% SVM classification accuracy when combining facial and eye-tracking features, compared to 66.67% with facial tracking alone and 64.58% with only eye-tracking.

Recent models integrating EEG data and facial/ocular metrics achieved 79% classification accuracy, with 76% sensitivity and 82% specificity (4). During COVID-19, Ghosh et al. used Twitter data for depression prediction, achieving 87.14% accuracy (5). Muzammel et al. developed an AI application using speech analysis, attaining 88% recognition precision (6). Worcester Polytechnic Institute’s AudiFace model, combining facial, audio, and textual data, underscored the importance of eye trajectory movements in diagnostics (7).

### Objective

Due to the lack of medical and economic resources, many developing regions struggle to provide adequate diagnosis and treatment for major depression (MD)(1). Current eye-tracking methods for assessing depression are complex and require costly equipment, limiting their clinical adoption. We have developed a non-wearable, cross-platform, cost-effective eye-tracking solution to address this issue using webcams. Our multimodal depression data collection system is affordable and scalable, enabling the creation of a comprehensive dataset that includes eye movement trajectories. By collaborating with our partner hospital, we are building a preliminary dataset. This system integrates various stimuli into a computerized interview, ensuring patient privacy while capturing instinctive responses for a thorough mental state assessment.

### Specific hypothesis

We hypothesize that major depression (MD) can be detected using computer vision strategies as patients may consciously conceal their symptoms. Studies show that combining machine learning with eye-tracking data is valuable for diagnosing MD.

## METHOD

### Participants

#### Inclusion criteria

Participants were selected from inpatients and outpatients with depression at the Fourth People’s Hospital of Hefei. The inclusion criteria are as follows:

► Clinical diagnosis of Major Depressive Disorder according to the Diagnostic and Statistical Manual of Mental Disorders (DSM-V), confirmed by clinical symptom evaluation by two attending psychiatrists or higher.
► Age 12 years and older.
► Willingness to participate in the study, with informed consent signed by the patient or the patient’s legal guardian.

#### Exclusion criteria

► Presence of severe physical illnesses, including inflammatory diseases and autoimmune diseases.
► Current or past neurological disorders or history of brain injury.
► Comorbid other psychiatric disorders or current psychotic symptoms.
► History of substance abuse.
► Currently undergoing electroconvulsive therapy (ECT) or transcranial magnetic stimulation (TMS).

### Stimuli and apparatus

The MDC system offers diverse and extendable stimuli options (Figure 1), all randomly triggered to eliminate interviewer bias, except for scales. Stimuli categories include:

**Figure 1.**
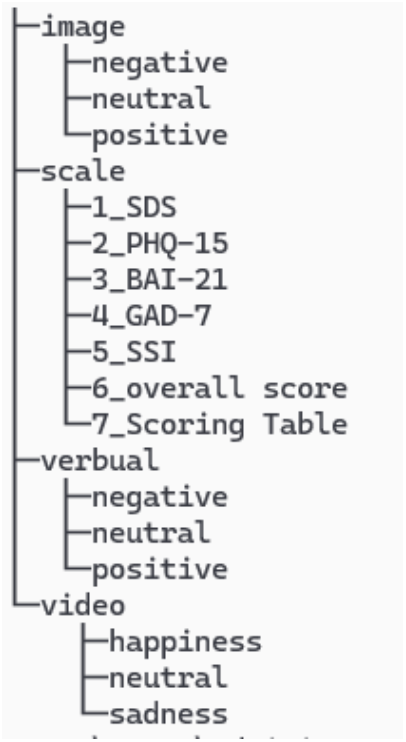
Stimuli file tree structure

1. Images: Divided into three sets (negative, neutral, positive), with 30 pre-selected images per set.
2. Scales: Five scales (SDS, PHQ-15, BAI-21, GAD-7, SSI) presented as multiple-choice questions, completed via monitor and mouse.
3. Text: 90 words (30 positive, 30 negative, 30 neutral) with pleasure, arousal, and frequency markers.
4. Videos: Three categories (comedy, tragedy, neutral), with one video from each category selected randomly.

#### Equipment and Setup

Our multimodal data collection system includes computers for doctors and patients, ergonomic chairs, detailed manuals, appropriate lighting, and wall-mounted spatial positioning scales. detailed items shown in Figure 3 while apparatus list is shown in Table 1.

**Table 1.** Apparatus and corresponding specifications.

| Item name | Model | Specification |
| --- | --- | --- |
| Interviewee's monitor | Xiaomi RMMNT27NF | •27 inches<br>•613.2*476.0mm active display area<br>•1920*1080 resolution |
| Interviewer's main monitor | Xiaomi RMMNT27NF | •27 inches<br>•613.2*476.0mm active display area<br>•1920*1080 resolution |
| Interviewer's secondary monitor | Lenovo B2213-2 | •21.5 inches<br>•508*392.9mm active display area<br>•1920*1080 resolution |
| In-person clinical assessment camera | Hikvision DS-E14a | 2k Resolution |
| Near-field Camera | Hikvision DS-D5ACAM100D | 1080p Resolution |
| Far-field Camera | The Hikvision DS-E12a | 4k Resolution |
| Spatial Location Camera | The Hikvision DS-E12a | 1080p Resolution |
| Light Bar | Xiaomi MJGJD01YL | •adjustable color temperature from 2700K to 6500K<br>•Default set to 4500K<br>•remote control enabled |
| Patient monitoring system | BerryMed PM6750 | ECG, Temperature, Spo2, HR and NIBP. we only enable HR and Spo2 functionalities. |

### Tasks

Tasks are controlled by our MDC software. Participants are positioned at a distance of 50-60cm from their respective screens. This deliberate spacing is intended to afford participants greater latitude in adjusting their seated posture, thereby enhancing their overall comfort during the session. The spatial location of each participant (location scale, Figure 2) is recoded by far field camera through entire data collection process.

**Figure 2.**
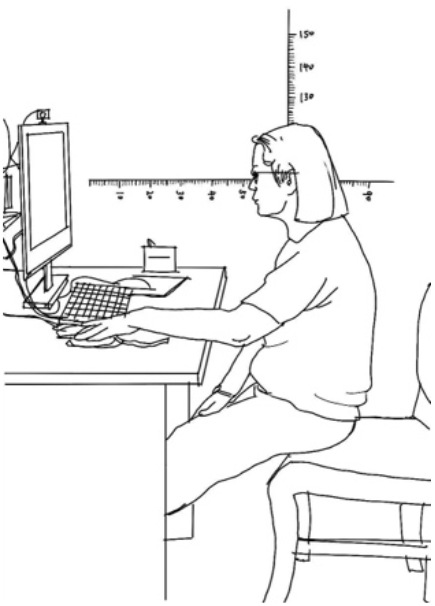
Participant’s spa2al loca2on. On the well, ver2cal and horizontal scales are implemented.

**Figure 3.**
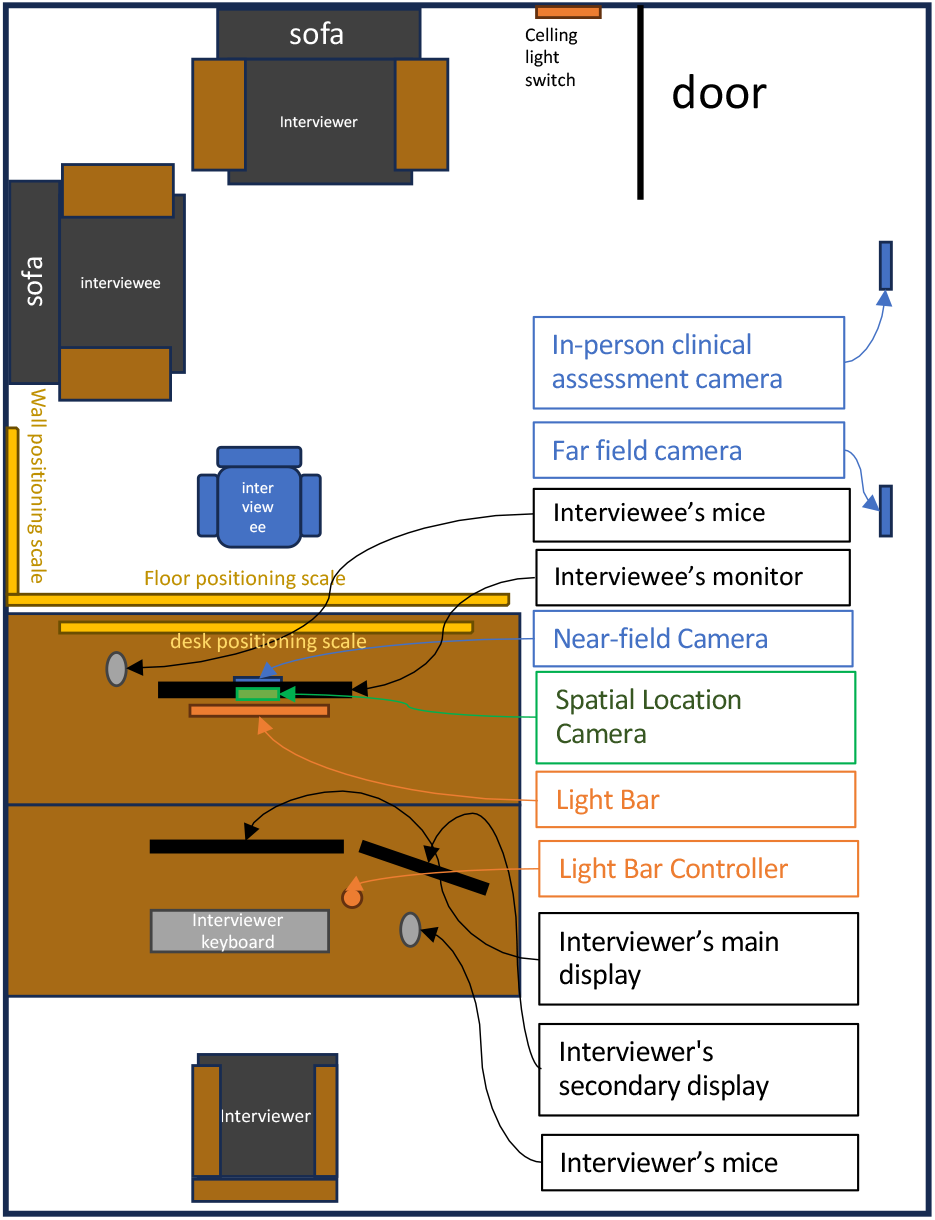
Sample of multimodal data collection room and the setup of all esseciental equipments

#### Multimodal Data Collection Tasks

The data collection session comprises several sub-sections, arranged chronologically: in-person clinical assessment, eye tracking calibration, depression scale task, image stimuli task, video stimuli task, and text stimuli task.

#### In-Person Clinical Assessment

The assessment session follows these steps:

1. The interviewer escorts the participant to the data collection room and ensures they are comfortably seated.
2. The interviewer checks the status of the ceiling light array.
3. The interviewer provides an overview of the data collection process and explains what to expect.
4. The interviewer presents the consent form, obtaining the participant’s signature if they agree.
5. A photocopy of the signed consent form is made for records.
6. The interviewer initiates phase 1 of the interview by starting the MDC’s graphical user interface (GUI), which activates the hidden far-field camera.

#### Eye Tracking Calibration

The MDC system activates all nearby cameras for the on-screen interview. A research assistant conducts a brief calibration using floor and wall positioning scales. Room lighting transitions from ceiling fixtures to an on-screen light bar. Hair pins are provided if necessary to prevent hair from obstructing the view of the ocular area. Participants are seated in the interviewer’s chair and align their head with a marked arrow sign. Calibration involves tracking a moving smiling face on the screen, ensuring precise gaze approximation. Comprehensive instructions are given verbally and visually (Figure 4).

**Figure 4.**
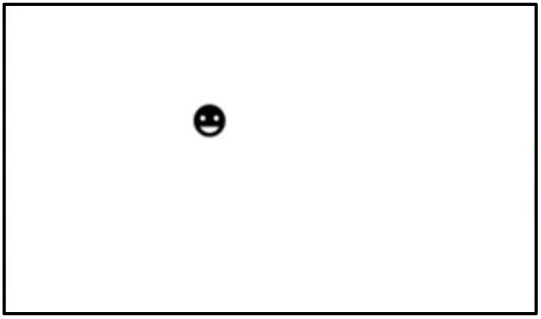
Sample of eye tracking calibration. Visual task.

#### Depression Scale Task

Post-calibration, participants engage in a series of sub-sections. The interviewer inputs scale results from the clinical assessment into the MDC database. The required scales are selected and sent to the participant’s screen. Participants interact with the digital scales using a mouse, with all scales presented in a multiple-choice text format. Cameras record 15 seconds before and after each interaction to capture emotional state changes.

#### Image, Video and Text Stimuli Task

Repeat scale task protocol for image, video and text stimuli sections. MDC software propagates automatically.

## RESULT

### Overview

Gaze-tracking technology provides an objective assessment of the visual content that draws attention from viewers of diverse backgrounds. We found that this type of data collection is especially helpful for modern machine learning strategies. We also learned about the video based gazing location extraction is achievable under our experiment setup. We are currently in the very early stage of our research work. We have obtained some preliminary findings in the areas of data collection enhancement, data security, and facial recognition. Further, we made micro adjustments in our experiment setup.

#### Session Duration and Complexity

The original on-screen assignments were tedious and comprised a total of 10 scales. Additionally, our preliminary run data were not utilized by medical professionals to identify changes in emotional state. To address these issues, we have revised the task schedule. Firstly, we condensed the scale task items from 10 to 5. As a result, the average time spent on the entire scales task decreased from 30 minutes to 9 minutes and 18 seconds. Furthermore, we introduced intervals between each item to better assess changes in emotional state (Table 2).

**Table 2.**
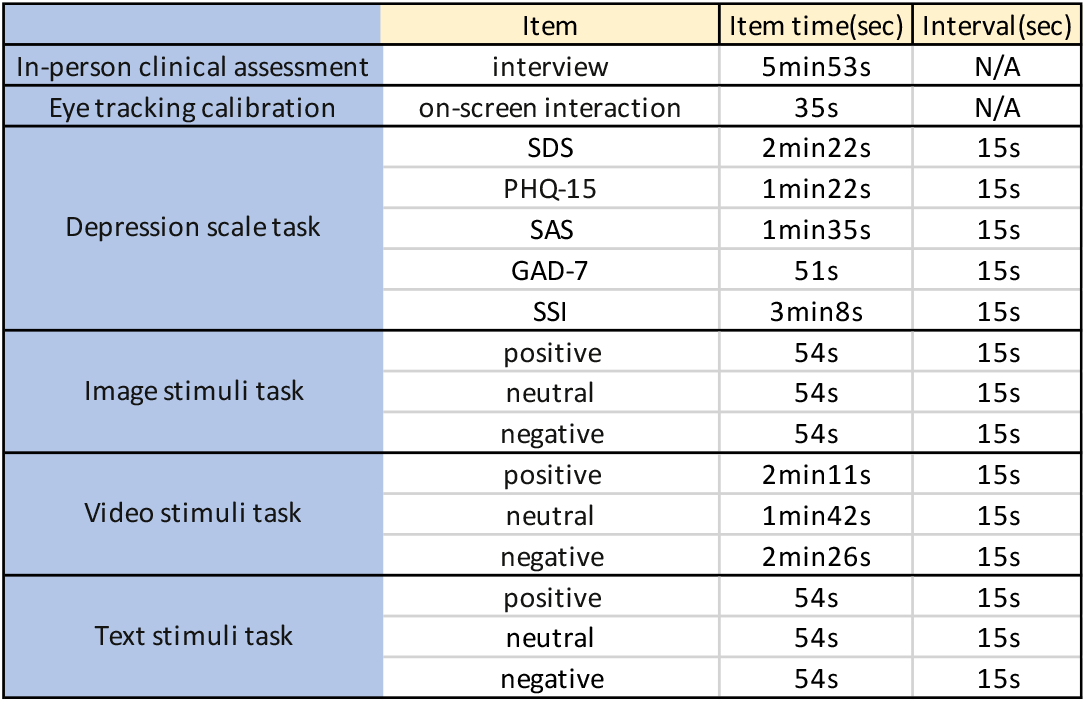
Timing table for MDC’s automatic session control.

|  | Item | Item time(sec) | Interval(sec) |
| --- | --- | --- | --- |
| In-person clinical assessment | interview | 5min53s | N/A |
| Eye tracking calibration | on-screen interaction | 35s | N/A |
| Depression scale task | SDS | 2min22s | 15s |
|  | PHQ-15 | 1min22s | 15s |
|  | SAS | 1min35s | 15s |
|  | GAD-7 | 51s | 15s |
|  | SSI | 3min8s | 15s |
| Image stimuli task | positive | 54s | 15s |
|  | neutral | 54s | 15s |
|  | negative | 54s | 15s |
| Video stimuli task | positive | 2min11s | 15s |
|  | neutral | 1min42s | 15s |
|  | negative | 2min26s | 15s |
| Text stimuli task | positive | 54s | 15s |
|  | neutral | 54s | 15s |
|  | negative | 54s | 15s |

#### Facial Data Extraction Result

After apply basic deep learning method, MTCNN, we are able to extract basic facial information for further machine learning research purpose. Other eye tracking methods such as Gaze360(8), Itracker(9), and our current research work will be implemented in the experiments for our future work.

By feeding in near field video to the convolutional neural network, we are able to extraction facial area image and approximating gaze direction.

## Discussion

### Principal Findings

This study demonstrates that high-quality data for computer science research can be gathered using webcams and other low-cost equipment. Eye gazing information can be extracted without using professional gaze-tracking tools. This scalable system, with appropriate machine learning networks, could significantly improve depression risk assessment in middle- and low-income countries

### Interpretation

During project creation, data and sessions are stored locally and can be transferred and merged into a larger project database. This approach enhances generalizability but also introduces challenges. After refining our algorithms, we plan to scale up MDC to be portable across various mobile devices and platforms, providing timely warnings to at-risk individuals.

## Data Availability

Non patient-identifiable data produced in the present study are available upon reasonable request to the authors

## Contributors

YFJ and HL were responsible for the design of the system architecture, the layout of the data collection room, and the development of the final study protocol. YD contributed valuable assistance in data collection execution. LXX oversees the maintenance and enhancement of software features throughout the duration of our study. JY, MH, and NA offer structural guidance for protocol development.

## Patient consent for publication

Not required

## Ethics approval

Ethical approval was provided by the National Health Commission of the PRC, Hefei Fourth People’s Hospital Ethics Committee (HSY-IRB-YJ-YYYX-JYF001). Results will be published in a peer-reviewed journal and presented at academic conferences.

## Provenance and peer review

Not commissioned; externally peer reviewed.

